# Point-of-care evaluation of a rapid antigen test (CLINITEST® Rapid COVID-19 Antigen Test) for diagnosis of SARS-CoV-2 infection in symptomatic and asymptomatic individuals

**DOI:** 10.1101/2021.02.02.21250984

**Authors:** Ignacio Torres, Sandrine Poujois, Eliseo Albert, Gabriela Álvarez, Javier Colomina, David Navarro

## Abstract

Rapid antigen assays (RAD) based on lateral flow immunochromatography (LFIC) technology have emerged as a valuable tool for the control of COVID-19 pandemic. Manufacturer□independent, real□world evaluation of these assays is crucial given the considerable heterogeneity reported in their clinical and analytical performances. Here, we report for the first time on the point-of-care performance of the CLINITEST® Rapid COVID-19 Antigen Test (Siemens, Healthineers, Erlangen, Germany) to detect SARS-CoV-2 infection in presumptive COVID-19 cases or asymptomatic close contacts of COVID-19 patients. When compared to RT-PCR, the overall sensitivity of the assay was 80.2 (95% CI, 70.9-87.1) for symptomatic patients sampled (nasopharyngeal specimens) within five days after the onset of symptoms and 60% (95% CI, 40.7-76.6%) for asymptomatic participants. The overall specificity was 100% in both population groups.

---

Rapid and decentralized testing for SARS-CoV-2 infection is one of the cornerstones of controlling the COVID-19 pandemic [1]. Rapid antigen assays (RAD) based on lateral flow immunochromatography (LFIC) technology offer advantages over molecular assays for the purpose, as they are low-cost, easy-to-use and instrument-free devices. An increasing number of RAD LFIC assays are being marketed nowadays. Manufacturer□independent, real□world evaluation of these assays is crucial given the considerable heterogeneity reported in their clinical and analytical performances [2]. Here, we report for the first time on the point-of-care (POC) performance of the CLINITEST® Rapid COVID-19 Antigen Test (Siemens, Healthineers, Erlangen, Germany) to detect SARS-CoV-2 infection in presumptive COVID-19 cases or asymptomatic close contacts of COVID-19 patients. The CLINITEST® RAD is a LFIC device licensed for detection of SARS-CoV-2 nucleocapsid protein in nasopharyngeal (NP) or nasal swabs to diagnose COVID-19 within the first week after symptoms onset.

A total of 270 subjects were enrolled in this prospective study from November 26 2020 to January 21 2021. The study was approved by the Ethical Committeee of Hospital Clínico Universitario INCLIVA, Valencia (October, 2021). Participants were either outpatients with suspected COVID-19 (n=178; median age, 41 years; range, 11-83 years; females, 112 -62.9%-), reporting at 5 or days or less (median 3 days; range 1-5 days) after onset of symptoms (one or more of the following: fever, dry cough, rhinorrhea, chest pain, dyspnea, myalgia, fatigue, anosmia, ageusia, odynophagia, diarrhea, conjunctivitis, and cephalea), or asymptomatic close contacts of COVID-19 patients (n=92; median age, 44 years; range, 11-87 years; females, 54 -58.7%-), as defined by the Spanish Ministry of Health [3]. Of the latter subset, 78 and 14 subjects were household or non-household contacts, respectively. These were sampled at a median of 4 days (range, 0-7 days) in the former group and a median of 5 days (range, 2-7 days) in the latter.

NP for RAD and RT-PCR testing were collected at POC by experienced nurses. For each patient, one swab was taken from the left nostril for RAD testing, and the other, obtained from the right nostril, was placed in 3 ml of Universal Transport Medium (UTM, Becton Dickinson, Sparks, MD, USA) and used for RT-PCR testing. RAD testing was carried out at POC immediately after sampling following the manufacturer’s recommendations. RT-PCRs were conducted within 24 h. of specimen collection at the Microbiology Service of Hospital Clínico Universitario (Valencia, Spain) with the TaqPath COVID-19 Combo Kit (ThermoFisher Scientific, Massachusetts, USA).

Among symptomatic patients, 73 tested positive by RT-PCR and RAD, 18 by RT-PCR only, and the remaining patients tested negative by both assays, thus indicating good agreement between RT-PCR and RAD results (Kappa index, 0,80; 95% CI, 0.71-0.88).

Household contacts (n=78; median age, 42 years; range, 14-81 years; females, 45 - 57.7% -) were tested at a median of 4 days (range, 0-7) after diagnosis of the index case. Non-household contacts (n=14; median age, 52 years; range, 11-87 years; range, 11-87 years; females, 9 -64.3%-) were sampled at a median of 5 days (range, 2-7) after self-reported exposure. Of the 15 subjects returning RT-PCR positive/RAD positive results, 10 tested positive only by RT-PCR, and the remaining 67 tested negative by both assays. The concordance between results returned by both assays was moderate in this population group (Kappa index, 0.68; 95% CI, 0.50-0.87). Interestingly, of the 15 asymptomatic participants testing positive by RT-PCR, five became symptomatic later on, all of whom had a positive RAD result.

As expected [2], SARS-CoV-2 RNA load was significantly higher (*P*<0.0001) in RT-PCR positive/RAD positive specimens than in RT-PCR positive/RAD negative samples from both symptomatic and asymptomatic participants (Supplementary Table 1).

**Table 1.** **Performance of the CLINITEST® Rapid COVID-19 Antigen Test at point of care for detection of SARS-CoV-2 infection**

| Population group | RAD performance |  |
| --- | --- | --- |
|  | Sensitivity (95% CI) | Specificity (95% CI) |
| Symptomatic patients with suspected COVID-19 | 80.2 (70.9-87.1) | 100% (95.8-100) |
| Asymptomatic close contacts of COVID-19 patients | 60.0 (40.7-76.6) | 100% (94.6-100) |
| All participants $C_T \leq 20$ ($\geq 7.1 \log_{10}$ copies/ml) | 100 (94.9-100) | - |
| All participants $C_T \leq 25$ ($\geq 6.2 \log_{10}$ copies/ml) | 95.5 (89-98.2) | - |
| All participants $C_T \leq 30$ ($\geq 5.2 \log_{10}$ copies/ml) | 94.6 (87.9-97.7) | - |
| All participants $C_T < 33$ ($\geq 3.4 \log_{10}$ copies/ml) | 75.9 (67.3-82.7) | - |
| CI, confidence interval; CT, RT-PCR cycle threshold; RAD, rapid antigen assay. |  |  |

Of note, time to sampling from symptoms onset or after exposure to the index case was similar across RAD-positive and RAD-negative participants (not shown).

Therefore, the sensitivity of the RAD assay was notably higher in symptomatic than in asymptomatic subjects (Table 1), whereas specificity was similar (100%). Negative and positive predictive values, adjusted to the median prevalence of positive cases within the study period in our Health Department (22%) were 94.7 % (95% CI, 87.6-97.9%) and 100 % (95% CI, 82.1-100%), respectively for symptomatic patients and 89.9% (95% CI, 81.7-94.6%) and 100% (95% CI, 77.5-100%), respectively for asymptomatic close contacts.

According to our data, the CLINITEST® Rapid COVID-19 Antigen Test meets the criteria recommended in WHO interim guidance for RAD diagnosis of SARS-CoV-2 infection (at least 80% sensitivity and 97% specificity), but as with other commercially-available RAD assays [1,5,6], this only applies in symptomatic patients with suspected COVID-19 who are tested shortly after symptoms onset (up to 5 days in the current study). In contrast, the POC performance of this and other RAD assays [1,6], is clearly suboptimal in asymptomatic close-contact individuals, either household or non-household. Two non-mutually exclusive factors may account for this observation: (i) SARS-CoV-2 RNA shedding in the upper respiratory tract (URT) could follow different kinetics in symptomatic and asymptomatic subjects, implying that the sampling time in the latter may have been inappropriate (too early or too late) to capture all infection cases; (ii) SARS-CoV-2 infected individuals not subsequently developing COVID-19 display lower overall viral loads in URT than those who do. In support of this latter viewpoint, all five participants eventually developing COVID-19 returned positive results by RAD at the sampling time. This was also observed in a previous study using the Panbio RAD assay from Abbott Laboratories [6].

## Data Availability

The data that support the findings of this study are available on request from the corresponding author.

## ACKNOWLEDGMENTS

We are grateful to Siemens Healthineers for providing the Rapid Test Device kits. This company had no role in the study design, data collection, data analysis, data interpretation, or writing of the report. We thank all staff working at Clinic University Hospital and primary healthcare centers belonging to the Clínico Malvarrosa Health Department for their unwavering commitment in the fight against COVID-19. We would also like to thank María José Beltrán, Pilar Botija and Ana Sanmartín for assistance in organizing RAD testing in primary healthcare centers.

## FINANCIAL SUPPORT

This work received no public or private funds.

## CONFLICTS OF INTEREST

The authors declare no conflicts of interest.

## AUTHOR CONTRIBUTIONS

IT, SP, EA and GA: Methodology and data validation. IT, EA, and JC: Formal analysis. DN: Conceptualization, supervision, writing the original draft. All authors reviewed the original draft.

## Notes

### Competing Interest Statement

The authors have declared no competing interest.

### Author Declarations

The study was approved by the Ethical Committeee of Hospital Clinico Universitario INCLIVA, Valencia (October, 2021)

