## Supplementary Table 1 for "Point-of-care evaluation of a rapid antigen test (CLINITEST® Rapid COVID-19 Antigen Test) for diagnosis of SARS-CoV-2 infection in symptomatic and asymptomatic individuals"

| **Supplementary Table 1. SARS-CoV-2 RNA load in nasopharyngeal specimens from patients testing either positive or negative by the CLINITEST® Rapid COVID-19 Antigen Test** | | |
| --- | --- | --- |
| Population group | Median C_T_ (range) | Median SARS-CoV-2 RNA load in log_10_ copies/ml (range)^a^ |
| Symptomatic patients with suspected COVID-19 testing RT-PCR positive/RAD positive | 15.0 (6.6-27.7) | 9 (5-11.7) |
| Symptomatic patients with suspected COVID-19 testing RT-PCR positive/RAD negative | 28.3 (21.2-32.8) | 4.8 (3.4-7.1) |
| Asymptomatic close contacts of COVID-19 patients testing RT-PCR positive/RAD positive | 15.9 (9.4-22.7) | 8.8 (6.6-10.8) |
| Asymptomatic close contacts of COVID-19 patients testing RT-PCR positive/RAD negative | 29.8 (23.8-32) | 4.4 (3.7-6.3) |
| C_T_, RT-PCR cycle threshold.  ^a^ The AMPLIRUN® TOTAL SARS-CoV-2 Control (Vircell S.A:, Granada, Spain) was used as the reference material for SARS-CoV-2 RNA load quantitation. | | |
